# Effectiveness of oral aciclovir in preventing maternal chickenpox: A comparison with VZIG

**DOI:** 10.1101/2022.04.26.22274015

**Authors:** Bersabeh Sile, Kevin E Brown, Charlotte Gower, Johanna Bosowski, Amanda Dennis, Michelle Falconer, Julia Stowe, Gayatri Amirthalingam

## Abstract

**Objectives:** Although often presenting as a self-limiting childhood disease, chickenpox can have serious consequences if acquired in pregnancy. The UK recommendations are that varicella immunoglobulin (VZIG) is administered intramuscularly to susceptible pregnant women exposed to chickenpox prior to 20 weeks gestation. Oral aciclovir or VZIG is recommended if exposure occurs at 20+ weeks gestation. Our objective was to compare the effectiveness of oral aciclovir to VZIG in preventing maternal and neonatal chickenpox.

**Methods:** We identified and followed up 186 pregnant women who were exposed to chickenpox and compared their outcomes.

**Results:** 171/186 (91.9%) of these women received either VZIG or oral aciclovir. 53 of the 145 (36.6%) women who received VZIG went on to develop chickenpox compared to 8 of the 26 (30.8%) women who received oral aciclovir (p=0.32). No statistical difference was found between the oral aciclovir and VZIG groups even after controlling for maternal age, gestational stage, type of exposure and IgG titre (adjusted OR:0.83; 95%CI:0.26-2.65; p=0.751).

**Conclusions:** These findings support the use of oral aciclovir as first-line prophylaxis in pregnant women exposed at 20+ weeks, (and possibly second-line <20 weeks) as they suggest its effectiveness at preventing maternal chickenpox is either better or equal to VZIG.

## Introduction

Chickenpox is a highly infectious disease caused by varicella-zoster virus (VZV). Illness is characterised by scattered vesicular skin lesions and fever and is usually self-limiting in otherwise healthy children. Chickenpox is found in a worldwide geographic distribution (1, 2) but is relatively more prevalent in temperate climates. Temperate countries without universal vaccination programmes can expect to have over 90% of their population infected by varicella by the age of 15 years (3), with incidence rates highest among children aged 1-4 years (3, 4). Primary varicella infection elicits the response of the protective immunoglobulin G (IgG) antibody (as well as IgM, and IgA antibodies) which confers life-long immunity. Migration into the UK from regions with lower VZV prevalence, combined with an estimated upward shift in the age distribution of chickenpox cases (5) has resulted in a sizable population of women of childbearing age who are VZV IgG sero-negative. These women are susceptible at a time in their lives where primary chickenpox infection poses a greater risk to them and their developing baby. Incidence of chickenpox among women of childbearing age is estimated to be at least around 2 - 3 /1,000 in the UK (6).

The most common complication of chickenpox in pregnancy is maternal pneumonia (7) which can be life threatening if not treated properly (8). Associated symptoms of cough, chest pain, fever, fatigue, and shortness of breath can further be exacerbated by symptoms of the later stages of pregnancy. Approximately 10% to 20% of pregnant women with acute chickenpox go on to develop pneumonia (9, 10).

Complications for the foetus and neonate include congenital varicella syndrome; a relatively rare condition which occurs in 0.39 % of maternal varicella cases in the first 20 weeks of infection (11). Affected new-borns may have low birth weight (intrauterine growth retardation), skin scar tissue on the arms and legs, and brain function abnormalities (9). The range and severity of symptoms vary greatly depending on when maternal infection occurred during foetal development. It is estimated around 1-2% of maternal varicella infections acquired in the first 20 weeks of pregnancy result in congenital varicella syndrome (9, 10). More than 40 cases of congenital varicella syndrome are estimated in the US each year with a mortality rate of approximately 30% in the first few months of life (9, 10).

Maternal varicella infection acquired in late pregnancy can result in the baby being born with neonatal varicella due to vertical transmission from mother to baby. Neonatal varicella is associated with a lower risk of long-term foetal damage than congenital varicella syndrome but severe and even fatal disease can occur especially if neonatal infection occurs within a week of birth (12).

Varicella vaccines (Varivax™ and Varilix ™) are live-attenuated vaccines and contraindicated in pregnancy, therefore, management of susceptible pregnant women is centred around trying to prevent or attenuate maternal chickenpox disease and associated complications by offering post exposure prophylaxis (PEP). Prior to August 2018, VZIG was recommended irrespective of gestational stage (14). However, due to a critical national shortage in July 2018, interim guidance was issued to restrict VZIG to women exposed in the first 20 weeks of pregnancy and neonates at risk of neonatal varicella infection in order to prioritise supply for those most at risk of severe complications (13). The decision to restrict oral aciclovir to women exposed after 20 weeks gestation was due to lack of evidence on its effectiveness at preventing congenital varicella syndrome, and the duration of antiviral treatment (13).

Although clinical studies have shown aciclovir to be effective at preventing or attenuating childhood chickenpox (15) real-world effectiveness data on aciclovir in pregnancy are extremely limited. The interim change in guidelines presented an opportunity for us obtain much needed on data on aciclovir effectiveness in pregnancy.

In this study we aim to compare the effectiveness of oral aciclovir to VZIG in preventing maternal and neonatal chickenpox disease using a prospective cohort study design.

## Patients and Methods

### Identification of Patients

Patients were identified via the Rabies and Immunoglobulin Service (RIgS) which is hosted by the Immunisation Division, UK Health Security Agency (UKHSA), Colindale. This service is run by a multi-disciplinary team of medical consultants, nurses and admin staff who support the post-exposure prophylaxis and clinical management of a range of infections including varicella. The RIgS handles requests from GPs, midwives and other clinicians who call in to request VZIG when presented with a pregnant woman exposed to varicella. The RIgS team will conduct a risk assessment, including requesting serum evidence of susceptibility (i.e. VZV IgG titres < 100 mlU/ml), and assessing the strength of exposure and then will either authorise the issuance of VZIG from one of the stockholders in the country or advise that aciclovir be prescribed locally. The type of prophylaxis recommended will follow the most current guidelines (13). Patients assessed by the RIgS team who fulfilled the following criteria were then contacted by our study team for their participation in the study.

The following inclusion criteria were used

- Patient had no history of chickenpox or a history of chickenpox vaccination
- Patient had an VZV IgG level of less than 100 mlU/ml
- Patient had more than 15 minutes face-face contact with a chickenpox case during the infectious period of the index case (48 hours prior to rash onset - 10 days after rash onset in index case)

### Data Collection

Patients identified as above were posted a questionnaire to their home address asking them to review the service they received, confirm the prophylaxis treatment they took and confirm whether they developed chickenpox. Demographic data such as date of birth and expected due date were also collected. Patients who did not return a completed postal questionnaire were subsequently contacted by telephone to collect this follow up information.

Once the expected due date had elapsed, the woman’s GP was contacted by the study team to ascertain the outcome of the birth and if there were any complications with the new-born.

Data obtained between Aug 2018 and Dec 2021 were analysed for this study. This study period was chosen to capture patients who were exposed after the change in the post-exposure management guidelines.

### Data Analysis

We compared the proportion of women in the aciclovir group who went on to develop chickenpox to that in the VZIG group. We also compared the proportion of women in the aciclovir group who went on to have possible VZV related complications in their new-born to that in the VZIG group. The Pearson’s chi-squared test was used to test the statistical significance of the difference between the two prophylaxis groups.

We fitted a multivariable logistic regression model to obtain the strength of association between each prophylaxis group and maternal chickenpox. The final model adjusted for VZV IgG level (< 50 mlU/ml vs 50+ mlU/ml), maternal age (< 30 years vs 30+ years), source of exposure (household contact vs non household) and gestational stage at exposure (< 20 weeks vs 20+ weeks). Data analyses were carried out in STATA 15™.

## Results

### Descriptive data of study population

There were 363 calls received by the RIgS team between 1^st^ August 2018 and 31^st^ December 2022 regarding susceptible pregnant women exposed to varicella infection. We obtained follow up data on 186 women who were assessed to have had significant exposure by the RIgS team and were deemed to be non-immune either qualitatively or quantitatively. 138/186 (74.2%) of these women were exposed before 20 weeks gestation, 42/186 (22.6%) of these women were exposed after 20 weeks gestation and 6/186 (3.2%) women did not have gestational stage recorded. The median maternal age was 34 years. The most common source of chickenpox exposure was the pregnant women’s older child with (165/186 women) 88.7% exposed by either their own child or another relative (e.g. niece or nephew) living within the same household.

### Univariate associations between prophylaxis group and maternal chickenpox

Of the 186 women we obtained data from, 171/186 (91.9%) were administered a post-exposure prophylactic treatment; 145/171 (84.8%) of which were administered VZIG and 26/171 (15.2%) took oral aciclovir (Figure 1). The 15/186 (8.1%) women who were in neither group were analysed separately.

**Figure 1.**
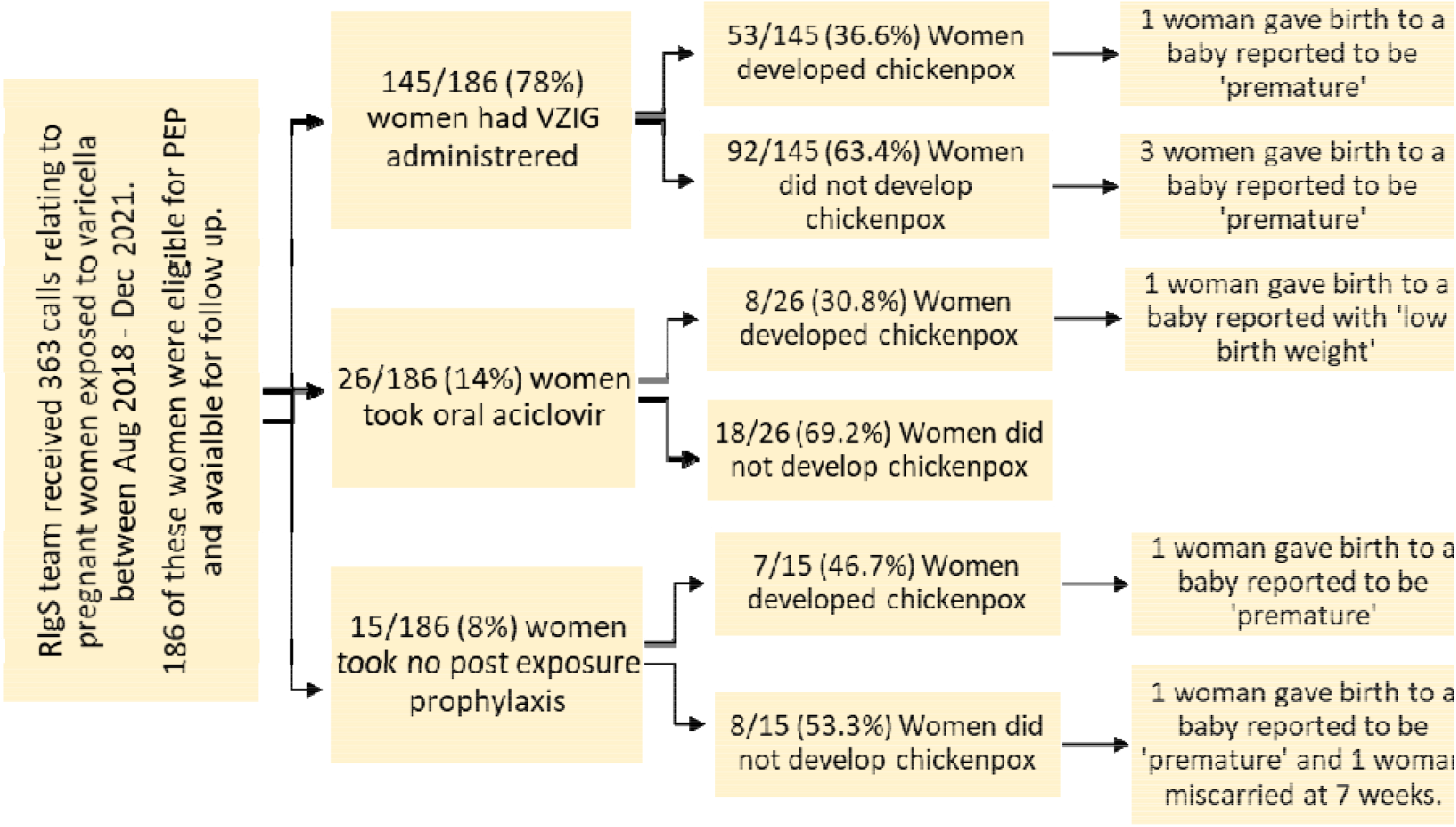
Study participants and their outcomes by PEP arm.

Of the 145 women who received VZIG 53/145 (36.6%) developed maternal chickenpox compared to 8/26 (30.8%) women who received aciclovir (Figure 1 and Table 1).

**Table 1.** Factors associated with maternal chickenpox. Results from the multivariable logistic regression.

|  | Number<br>(row %) | Adjusted Odds<br>Ratio | 95 % Confidence<br>Interval | p-value |
| --- | --- | --- | --- | --- |
| **Type of PEP** |  |  |  |  |
| VZIG | 53 (36.6%) | 1 (reference) |  |  |
| Aciclovir | 18 (30.8%) | 0.83 | 0.26 - 2.60 | 0.751 |
| No PEP | 7 (46.7%) | 1.54 | 0.32 - 7.48 | 0.595 |
| **Gestational Stage** |  |  |  |  |
| < 20 weeks | 51 (37.0%) | 1 (reference) |  |  |
| 20+ weeks | 13 (31.0%) | 0.68 | 0.28 - 1.66 | 0.400 |
| **VZV IgG Level** |  |  |  |  |
| 0 – 50 (mIU/ml) | 47 (43.1%) | 1 (reference) |  |  |
| 50 – 100 (mIU/ml) | 8 (15.1%) | 0.18 | 0.077- 0.44 | < 0.05 |
| **Source of Varicella Exposure** |  |  |  |  |
| Relative from same household (i.e. own child/niece/nephew) | 64 (38.8%) | 1 (reference) |  |  |
| Child in the school where the woman works/Child in same hospital waiting room | 2 (10.5%) | 0.12 | 0.025 - 0.56 | 0.007 |
| **Maternal Age** |  |  |  |  |
| < 30 years | 15 (35.7%) | 1 (reference) |  |  |
| 30 + years | 53 (37.1%) | 1.23 | 0.57 - 2.69 | 0.595 |
| PEP - Post Exposure Prophylaxis |  |  |  |  |
| VZIG - Human Varicella Immunoglobulin |  |  |  |  |
| VZV - Varicella Zoster Virus |  |  |  |  |

The difference in the two proportions was not statistically significant. (Pearson’s chi squared test. p-value = 0.3213). Just over half of the women who developed maternal chickenpox in the VZIG group reported having more than 30 lesions, compared to 25% in the aciclovir group (Table 2).

**Table 2.** Severity of maternal chickenpox by prophylaxis group.

|  | Number of Chickenpox Lesions |  |  |  | Total |
| --- | --- | --- | --- | --- | --- |
|  | Less than 30 | Ten to 30 | More than 30 | Not Known |  |
| Aciclovir | 3 (37.5%) | 3 (37.5%) | 2 (25.0%) | 0 | **8** |
| VZIG | 8 (15.1%) | 14 (26.4%) | 27 (50.9%) | 4 (7.5%) | **53** |
| No PEP | 0 | 3 (42.9%) | 0 | 4 (57.1%) | **7** |
| **Total** | **11** | **20** | **29** | **8** | **68** |

4/145 (3%) women from the VZIG group gave birth to a child who was either premature or had a low birth weight and 1/26 (4%) woman in the ACV group gave birth to a child with low birth weight (Figure 1). 14/145 (10%) women in the VZIG group reported pain at site of injection. None of the women in the aciclovir group reported side effects.

7/15 (46.7%) of the women who received no prophylaxis went on to develop maternal chickenpox.

### Multivariable associations between prophylaxis group and maternal chickenpox

After controlling for maternal age, gestational stage, IgG level and source of exposure the odds of developing maternal chickenpox in the aciclovir group was lower than for the VZIG group (Table 1). However, this difference was not found to be statistically significant (adjusted OR: 0.83, 95%CI 0.26 – 2.65; p=0.751) (Table 1).

The odds of developing maternal chickenpox was significantly lower in women exposed to a person outside of their household (e.g. hospital waiting room, child in nursery where pregnant woman is a teacher or playdate with friend’s child) (adjusted OR: 0.12; 95%CI 0.025 – 0.56; p=0.007) (Table 1). As was the odds for women with an IgG level of 50+ mlU/ml (adjusted OR: 0.18; 95%CI 0.079 – 0.37; p<0.05) (Table 1). Maternal age and gestational stage were not found to be significantly associated with maternal chickenpox (Table 1).

## Discussion

We found no statistical difference between the effectiveness of oral aciclovir and VZIG in preventing maternal chickenpox. A lower proportion of women who were given aciclovir developed chickenpox compared to those given VZIG (30.8% vs 36.6%) but this difference was not statistically significant. These findings suggest that oral aciclovir is at least as effective as VZIG at preventing maternal chickenpox following varicella exposure.

Our findings are supported by results from data collected in 2019/2020 from a sentinel network of virologists. These data showed favourable results for the effectiveness of aciclovir in preventing maternal chickenpox. None of the 12 successfully followed up pregnant women who received oral aciclovir post varicella exposure went on to develop maternal chickenpox compared to 1/9 pregnant woman (11%) who developed maternal chickenpox after receiving VZIG (unpublished data).

### Strengths and Limitations

One of the biggest strengths of our study is that it is the first to examine the effectiveness of oral aciclovir in pregnant women in a ‘real word’ setting. Although clinical trials have demonstrated its efficacy there is very little data on the effectiveness of aciclovir in pregnancy, and no data on the effectiveness of preventing congenital varicella infection.

Another strength is the robustness of the data source used. Data from the RIgS is a good data source for identifying women eligible for VZIG. VZIG is now stored at only a limited number of stockholders with the approval to issue being largely centralised. Furthermore, due to the risk assessment required to issue VZIG we were able to capture data on, and control for, other determinants of infection such as VZV IgG levels and type of exposure.

Conversely to clinicians having to request VZIG, clinicians prescribing aciclovir do not need to contact RIgS as this can be prescribed locally. Therefore, a major limitation of our study is that we are unlikely to have captured the majority of women prescribed aciclovir. The relatively small number of pregnant women recruited into the aciclovir group coupled with the rarity of varicella associated new-born outcomes meant that we were unable to fully assess the effectiveness of aciclovir on preventing varicella related complications in the new-born.

### Other countries who recommend oral aciclovir

The UK is not the only country to recommend oral aciclovir as post exposure prophylaxis. Oral aciclovir is used as a second line option in Australia and New Zealand where VZIG is recommend as first-line if the pregnant woman presents within 4 days of exposure and then oral aciclovir is recommended if she presents after that time. They recommend restricting aciclovir to women exposed in the second half of pregnancy unless the woman is a smoker, is immunocompromised or has underlying lung disease, in which case oral aciclovir can be given prior to 20 weeks gestation in these instances (16-18).

### Concluding remarks

Assessing the effectiveness of oral aciclovir prophylaxis is important as it could serve as a useful and possibly more effective alternative to VZIG, especially in situations where VZIG is unavailable or cannot be given. Our findings support the recommendation of oral aciclovir being given as first line prophylactic therapy in pregnant women exposed at 20+ weeks gestation. Further work is required to ascertain whether women exposed before 20 weeks could also benefit from oral aciclovir prophylaxis. There is evidence to suggest that if guidelines changed to recommend aciclovir to women prior to 20 weeks gestation then this would be safe to do so (19).

## Data Availability

All data produced in the present work are contained in the manuscript

## Ethics

The review for the study was conducted by the Public Health England (PHE) Research Ethics and Governance of Public Health Practice Group (REGG). The review covered the study design, content and feasibility, and all legal, financial, regulatory and ethical considerations. As a result of this review, the study was categorized as a service evaluation. As no ethical issues were identified it was decided that consideration by an ethics committee would not be necessary.

## Acknowledgements

The authors thank the team at the Rabies and Immunologlobulin Service (RIgS) (with special thanks to Ms Teresa Gibbs, Ms Deborah Cohen and Ms Donna Haskins) for their logistical support throughout the study.

